# Echocardiographic Characteristics, Measures of Severity and Natural History of Isolated Calcific Mitral Stenosis

**DOI:** 10.64898/2026.05.11.26352948

**Authors:** Jeremiah Haines, Tyler Jacobson, Solomon Ocran, Lindsey Kalvin, Victor Redmon, Liyun Zheng, Amy Pan, Noelle Garster, David Lewandowski, Michael Widlansky, Divyanshu Mohananey

**Affiliations:** Division of Cardiovascular Medicine, Department of Medicine, Medical College of Wisconsin, Milwaukee, USA; Division of Internal Medicine, Department of Medicine, Medical College of Wisconsin, Milwaukee, USA; Quantitative Health Sciences, Department of Pediatrics, Medical College of Wisconsin, Milwaukee, USA

**Keywords:** Calcific mitral stenosis, mitral annular calcification, mitral valve replacement

## Abstract

**Introduction:** With improved life expectancy, mitral annular calcification and calcific mitral stenosis (CMS) are increasing in prevalence. Echocardiographic evaluation of CMS is challenging due to acoustic shadowing and lack of CMS specific data on assessment of severity and outcomes.

**Methods:** We retrospectively identified patients with isolated CMS between the years 1/1/2010 and 4/5/2022. Severe CMS was defined as MVA_cont_ ≤1.5 cm^2^. The primary outcome was a composite of all-cause mortality, mitral valve replacement (MVR) and ischemic stroke. Outcomes were collected through electronic health records with follow up through 8/15/2025.

**Results:** Our cohort included a total of n=717 patients with CMS of which n=140 had severe CMS. The mean age was 74±13 years and cohort was predominantly female. We found that MVA_PHT_ consistently overestimates the MVA and is a poor predictor of severe CMS. Mean gradient >5 mm Hg had 81% specificity and 57% sensitivity for severe CMS. Over a median follow up of 36 (IQR 10.5-49.7) months, a total of n=331 (46.2%) patients died, and the primary composite outcome occurred in n=370 (51.6%). Although MVA_cont_ ≤1.5 cm^2^ [aHR 1.3 (95% CI 0.9-1.8),p=0.29] was not an independent predictor of the primary outcome we found that mTMG was a significant independent predictor primary outcome [aHR 1.5 (95% CI 1.1-2), p<0.01]. Patients with MVA_cont_ ≤1.5 cm^2^ and mean gradient ≥ 5 mmHg had the highest risk for the primary outcome [aHR 2 (95% CI 1.1-3.7),p=.02].

**Conclusion:** Patients with severe CMS are older, female with a high burden of comorbidities and carry an overall poor prognosis. mTMG is an independent prognostic marker in these patients. Patients with MVA ≤1.5 cm^2^ and mTMG ≥5 mmHg have the worst prognosis.

## INTRODUCTION

With significant improvements in global living standards, greater access to healthcare and availability of antimicrobials, the prevalence of rheumatic heart disease has markedly declined in developed nations.^1^ Conversely, with improved life expectancy, the prevalence of degenerative valvular diseases including mitral annular calcification (MAC) and calcific mitral stenosis (CMS) has increased in the aging population. MAC has been reported in 23% of patients referred for echocardiography at Mayo Clinic and in up to 42% of community dwelling elderly persons. ^2,3^ Despite its rising prevalence, assessment of patients with CMS remains challenging due to limitations of echocardiographic evaluation and lack of CMS specific data. We have previously outlined these challenges using case-based examples from our echocardiography lab. ^4^ Traditional markers of severity in rhematic mitral stenosis such as pressure half time (PHT) and 2D planimetry are frequently unreliable or challenging to measure due to the acoustic shadowing from the calcified annulus and leaflets. Mean transmitral gradient (mTMG), although easy to perform, can be both *falsely elevated* due to high filling pressures, diastolic dysfunction and poor left atrial compliance or *falsely reduced* due to lower stroke volumes in these elderly individuals with smaller cavities and concentric hypertrophy.^5–8^ Despite these limitations, limited literature suggests that mTMG retains prognostic value in CMS. ^9 10^ Currently, continuity equation derived mitral valve area (MVA_cont_) is considered the reference standard for evaluation of CMS despite challenges with measurement and limited prognostic data.^11^ To address key gaps literature, we aimed to 1) characterize echocardiographic and clinical features of patients with CMS, 2) evaluate the diagnostic performance of PHT and mTMG with increasing severity of CMS 3) assess clinical outcomes of patients with CMS and the independent prognostic value of MVA_cont_ and mTMG.

## MATERIALS AND METHODS

Patient population: We retrospectively identified patients in our echocardiographic dataset who underwent transthoracic echocardiograms (TTE) between the years 1/1/2010 and 4/5/2022 and had a mTMG ≥ 2 mmHg. The study was approved by the local institutional review board. Inclusion criteria included any patient with MAC and elevated TMG (≥ 2 mmHg). Exclusion criteria included patients with rheumatic mitral valve disease, moderate or severe mitral regurgitation, moderate or severe aortic regurgitation, prior mitral valve surgery (repair or replacement), native or prosthetic aortic valve area <1.5 cm2, poor quality Doppler measurements or inability to measure the left ventricular outflow tract diameter (LVOTd). All studies were manually reviewed to ensure accurate identification of exclusion criteria (supplementary appendix).

### Echocardiography

All measurements were performed according to the American Society of Echocardiography’s standardized reporting guidelines. An average of 3 cardiac cycles was used for all Doppler measurements. Each included echocardiogram was manually reviewed independently by either a level 3 echocardiographer (NG, DM, DL) or experienced echocardiography research core lab sonographer (LK) to ensure accuracy of measurements and inclusion of any missing data. Discrepancies were adjudicated by consensus. Extent of MAC was assessed in the parasternal short-axis view and was noted to be circumferential if 360 degrees of MAC were visualized. Leaflet calcification as well as sub-valvular calcification was assessed on the apical 4 chamber and 3 chamber views. Mitral valve area by continuity (MVA_cont_) was derived using the formula: *π x* (*LVOTd*/2)2 *x LVVTI*/*MVVTI* and MVA_PHT_ was derived using the formula: *220/PHT*. Mild CMS was defined as MVA>2cm^2^, moderate CMS as MVA 1.6-2 cm^2^ and severe CMS as ≤ 1.5cm^2^. Lastly, we conducted an independent interobserver variability assessment for the LVOTd and Doppler measurements for 10% of the sample. For LVOTd, we found the intraclass correlation coefficient (ICC) demonstrated moderate agreement between measurements (ICC 0.59, 95% CI 0.38–0.73, p < 0.001). For deceleration time, we found moderate agreement (ICC 0.51, 95% CI 0.30–0.67, p < 0.001). For MV VTI (ICC 0.92, 95% CI 0.86–0.96, p < 0.001) and mTMG (ICC 0.94, 95% CI 0.91–0.97, p < 0.001) there was high interobserver agreement.

#### Baseline Clinical Characteristics

Baseline clinical characteristics included age, sex, history of coronary artery disease (CAD), prior history of percutaneous intervention percutaneous intervention, prior history of coronary artery bypass grafting, prior history of myocardial infarction, diabetes, hypertension, chronic kidney disease (CKD), history of end-stage renal disease on dialysis (ESRD), prior stroke, atrial fibrillation, congestive heart failure (CHF), dyslipidemia, cirrhosis and peripheral arterial disease (PAD). These comorbidities were identified using ICD codes and then verified by manual chart review.

#### Outcomes

The Primary outcome was a composite of all-cause mortality, mitral valve replacement (MVR) and ischemic stroke. The secondary outcome was a composite of all cause mortality and MVR. Outcomes were collected through electronic health records with follow up through 8/15/2025.

### Statistical Analysis

Data were assessed for normal distribution using the Shapiro Wilk test. Categorical variables are expressed as percentages and continuous variables as mean±SD or median (interquartile range) for skewed variables. We used the Pearson χ2 test for univariate analysis of categorical variables and the independent samples test for univariate analysis of continuous variables. For non-parametric continuous variables, the Man-Whitney U test was used for univariate analysis. We conduced receiver operator curve (ROC) analyses to study the utility of PHT and mean TMG in predicting an MVA ≤ 1.5 cm^2^. Un-adjusted and adjusted Cox-Regression analyses were performed to assess the association of MVA and mean TMG with the primary and secondary outcomes. Cox-regression models were adjusted for age, sex, co-morbidities (CAD, diabetes, CKD, ESRD, prior stroke, atrial fibrillation, CHF, dyslipidemia, cirrhosis and PAD), left atrial size, left ventricular size (left ventricular end diastolic volume), left ventricular ejection fraction (LVEF), right ventricular size, right ventricular function. Right ventricular systolic pressure (derived from tricuspid regurgitation maximum velocity) was not added to the model due to a high percentage of missing values (31%). A 2-tailed *P*<0.05 was used to denote statistical significance. Statistical analyses were performed using SAS 9.4 (SAS Institute Inc., Cary, NC) and IBM SPSS Statistics (Version 28.0, Armonk, NY).

## RESULTS

Our cohort included a total of n=717 patients with CMS of which n=140 had severe CMS. The mean age was 74±13 years and cohort was predominantly female. There was a high burden of comorbidities with 67% patients having CAD, 62% having diabetes, 80% having either ESRD or CKD, 44% having a history of AF and 65% having CHF (Table 1). Patients with severe CMS were older, and more likely to be female. Prevalence of comorbidities was similar between mild, moderate and severe CMS except for prior stroke and PAD which were more prevalent in patients with severe CMS.

**Table 1:** Demographics, Co-morbidities and Echocardiographic characteristics*.

|  | Overall Cohort (n=717) | Mild CMS (MVA >2cm<sup>2</sup>) (n=333) | Moderate CMS (MVA 1.6-2 cm<sup>2</sup>) (n=244) | Severe CMS (<=1.5cm<sup>2</sup>) (n=140) | p-value |
| --- | --- | --- | --- | --- | --- |
| **Demographics** |  |  |  |  |  |
| Age | 76.0 (66.0-84.0) | 74.0 (65.0-82.0) | 77.0 (67.0-85.0) | 78.0 (68.5-86.0) | 0.002 |
| Female | 550 (69.7) | 214 (64.3) | 183 (75) | 103 (73.6) | 0.012 |
| **Co-morbidities** |  |  |  |  |  |
| Coronary artery disease | 480 (67)) | 216 (64.9) | 169 (69.3) | 95 (67.9) | 0.52 |
| History of prior PCI | 66 (9.2) | 26 (7.8) | 28 (11.5) | 12 (8.6) | 0.31 |
| History of prior CABG | 55 (7.7) | 29 (8.7) | 16 (6.6) | 10 (7.1) | 0.61 |
| History of prior myocardial infarction | 179 (25) | 73 (22) | 69 (28.3) | 37 (26.4) | 0.20 |
| Diabetes | 442 (61.7) | 202 (60.7) | 149 (61.1) | 91 (65.0) | 0.66 |
| Hypertension | 620 (86.5) | 283 (85.0) | 231 (87.3) | 124 (88.6) | 0.52 |
| Chronic Kidney Disease | 416 (58) | 192 (57.7) | 139 (57) | 85 (60.7) | 0.76 |
| ESRD on HD | 159 (22.2) | 82 (24.6) | 49 (20.1) | 28 (20.0) | 0.34 |
| Prior Stroke | 206 (28.7) | 81 (24.3) | 78 (32) | 47 (33.6)<sup>a</sup> | 0.050 |
| Atrial Fibrillation | 317 (44.2) | 138 (41.4) | 110 (45.1) | 69 (49.3) | 0.28 |
| Congestive Heart Failure | 466 (65.0) | 219 (65.8) | 153 (62.7) | 94 (67.1) | 0.63 |
| Prior Aortic Valve Replacement | 15 (2.1) | 7 (2.1) | 3 (1.2) | 5 (3.6) | 0.29 |
| Dyslipidemia | 597 (83.3) | 281 (84.4) | 203 (83.2) | 113 (80.7) | 0.62 |
| Cirrhosis | 63 (8.8) | 35 (10.5) | 16 (6.6) | 12 (8.6) | 0.25 |
| Peripheral arterial disease | 224 (31.2) | 92 (27.6) | 79 (32.4) | 53 (37.9)<sup>a</sup> | 0.08 |

\*Data were presented as n (%) or median (IQR)
Abbreviations: PCI: percutaneous intervention, CABG coronary artery bypass grafting, ESRD: end stage renal disease, HD: hemodialysis, MVA: mitral valve area, CMS: calcific mitral valve stenosis
<sup>a</sup> Although overall p-values was non-significant, between group differences (severe mitral stenosis vs. Mild mitral stenosis were significantly different, p <0.05)

### Echocardiographic characteristics

#### MAC and leaflet calcification

Circumferential MAC was present in only 4.5% of patients. Posterior leaflet involvement was more frequent than anterior leaflet involvement. We also found that 29% of patients had MAC with elevated mean TMG without any leaflet calcification. Involvement of the sub-valvular structures (chordae/papillary muscles) was seen in up to 1/3^rd^ of the cohort. Patients with severe CMS were more likely to have circumferential MAC, bi-leaflet calcification, and sub-valvular calcification (Table 2).

**Table 2:**
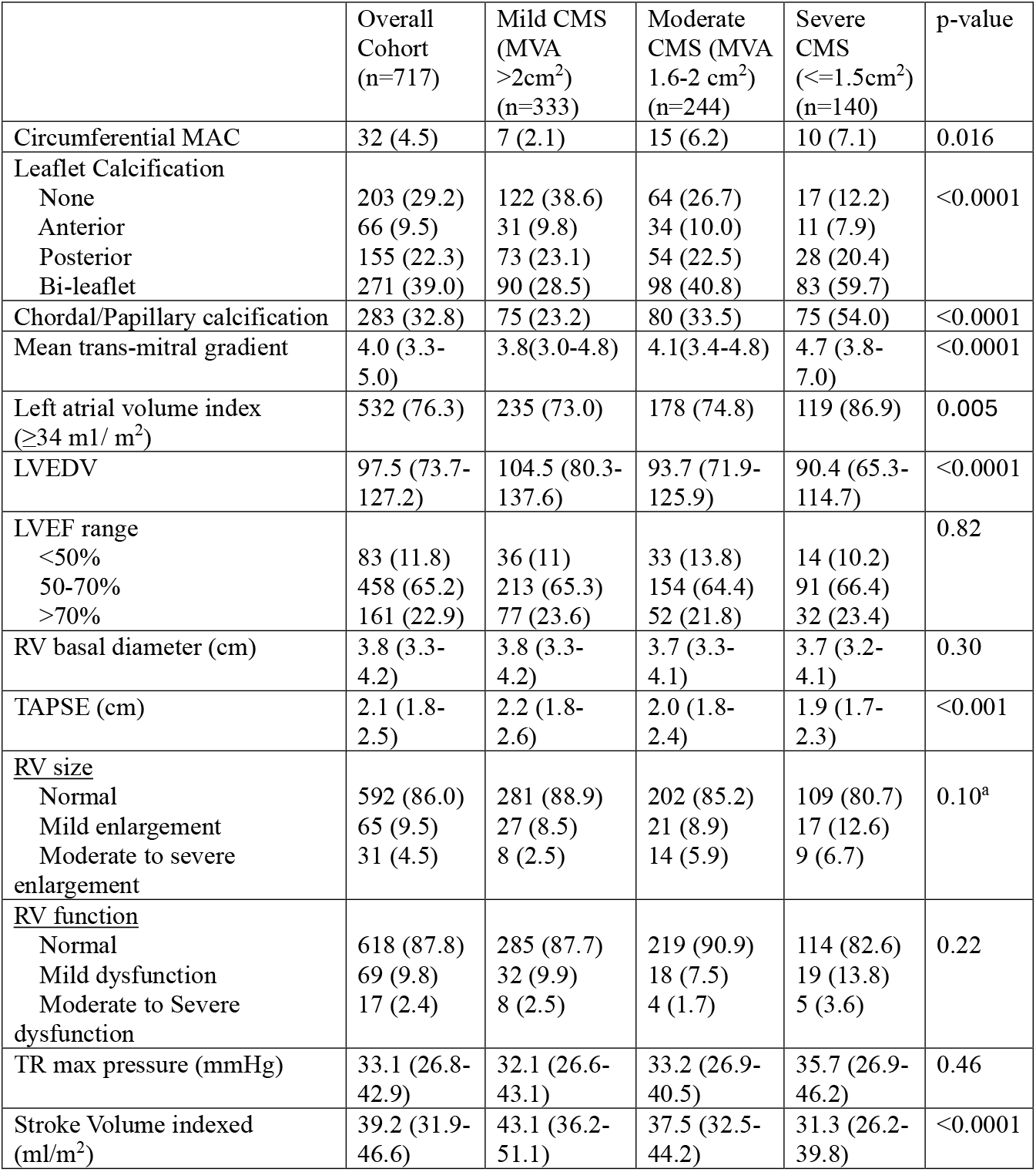
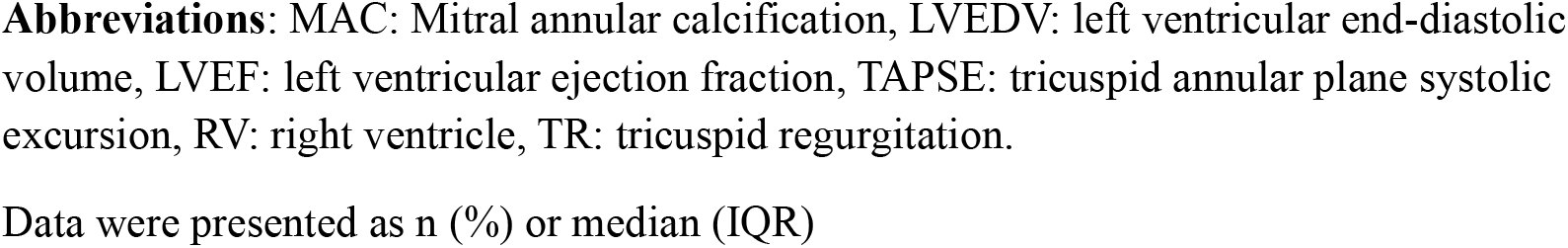
Echocardiographic characteristics.

|  | Overall Cohort (n=717) | Mild CMS (MVA >2cm<sup>2</sup>) (n=333) | Moderate CMS (MVA 1.6-2 cm<sup>2</sup>) (n=244) | Severe CMS (<=1.5cm<sup>2</sup>) (n=140) | p-value |
| --- | --- | --- | --- | --- | --- |
| Circumferential MAC | 32 (4.5) | 7 (2.1) | 15 (6.2) | 10 (7.1) | 0.016 |
| Leaflet Calcification |  |  |  |  | <0.0001 |
| None | 203 (29.2) | 122 (38.6) | 64 (26.7) | 17 (12.2) |  |
| Anterior | 66 (9.5) | 31 (9.8) | 34 (10.0) | 11 (7.9) |  |
| Posterior | 155 (22.3) | 73 (23.1) | 54 (22.5) | 28 (20.4) |  |
| Bi-leaflet | 271 (39.0) | 90 (28.5) | 98 (40.8) | 83 (59.7) |  |
| Chordal/Papillary calcification | 283 (32.8) | 75 (23.2) | 80 (33.5) | 75 (54.0) | <0.0001 |
| Mean trans-mitral gradient | 4.0 (3.3-5.0) | 3.8(3.0-4.8) | 4.1(3.4-4.8) | 4.7 (3.8-7.0) | <0.0001 |
| Left atrial volume index (≥34 ml/ m<sup>2</sup>) | 532 (76.3) | 235 (73.0) | 178 (74.8) | 119 (86.9) | 0.005 |
| LVEDV | 97.5 (73.7-127.2) | 104.5 (80.3-137.6) | 93.7 (71.9-125.9) | 90.4 (65.3-114.7) | <0.0001 |
| LVEF range |  |  |  |  | 0.82 |
| <50% | 83 (11.8) | 36 (11) | 33 (13.8) | 14 (10.2) |  |
| 50-70% | 458 (65.2) | 213 (65.3) | 154 (64.4) | 91 (66.4) |  |
| >70% | 161 (22.9) | 77 (23.6) | 52 (21.8) | 32 (23.4) |  |
| RV basal diameter (cm) | 3.8 (3.3-4.2) | 3.8 (3.3-4.2) | 3.7 (3.3-4.1) | 3.7 (3.2-4.1) | 0.30 |
| TAPSE (cm) | 2.1 (1.8-2.5) | 2.2 (1.8-2.6) | 2.0 (1.8-2.4) | 1.9 (1.7-2.3) | <0.001 |
| <u>RV size</u> |  |  |  |  | 0.10<sup>a</sup> |
| Normal | 592 (86.0) | 281 (88.9) | 202 (85.2) | 109 (80.7) |  |
| Mild enlargement | 65 (9.5) | 27 (8.5) | 21 (8.9) | 17 (12.6) |  |
| Moderate to severe enlargement | 31 (4.5) | 8 (2.5) | 14 (5.9) | 9 (6.7) |  |
| <u>RV function</u> |  |  |  |  | 0.22 |
| Normal | 618 (87.8) | 285 (87.7) | 219 (90.9) | 114 (82.6) |  |
| Mild dysfunction | 69 (9.8) | 32 (9.9) | 18 (7.5) | 19 (13.8) |  |
| Moderate to Severe dysfunction | 17 (2.4) | 8 (2.5) | 4 (1.7) | 5 (3.6) |  |
| TR max pressure (mmHg) | 33.1 (26.8-42.9) | 32.1 (26.6-43.1) | 33.2 (26.9-40.5) | 35.7 (26.9-46.2) | 0.46 |
| Stroke Volume indexed (ml/m<sup>2</sup>) | 39.2 (31.9-46.6) | 43.1 (36.2-51.1) | 37.5 (32.5-44.2) | 31.3 (26.2-39.8) | <0.0001 |

**Abbreviations:** MAC: Mitral annular calcification, LVEDV: left ventricular end-diastolic volume, LVEF: left ventricular ejection fraction, TAPSE: tricuspid annular plane systolic excursion, RV: right ventricle, TR: tricuspid regurgitation.
Data were presented as n (%) or median (IQR)

#### Atrial volumes, ventricular volumes and function

Left atrial enlargement [indexed left atrial volume (LAVi) ≥ 34 ml/m^2^] was present in 76% of the cohort, with a significantly higher prevalence in patients with severe CMS. LVEF was preserved in 88% of the cohort with no differences between the groups. We also found that patients with severe CMS had significantly smaller left ventricular sizes. Majority of the cohort had normal right ventricular (RV) size and function. Although the overall between group differences were not significant for RV size, we did find that patients with severe CMS were more likely to have RV enlargement than those with mild CMS. Though average TAPSE was lower in patients with greater severity of CMS, echocardiographer assessed multiparametric assessment of RV function showed no difference between groups (Table 2).

#### PHT, mean gradient and MVA_cont_

We found that MVA_PHT_ consistently overestimates the MVA. Of the 228 patients with moderate CMS by MVA_cont,_ 70% were classified as mild stenosis by MVA_PHT_. Similarly, of the n=133 patients with severe CMS by MVA_cont_, 68% were classified as mild and 22% were classified as moderate stenosis by MVA_PHT_ (Figure 1). We further conducted an ROC analysis to study the ability of PHT to predict severe CMS and found it to be a poor predictor with an area under curve (AUC) of 0.54 (supplementary appendix).

**Figure 1:**
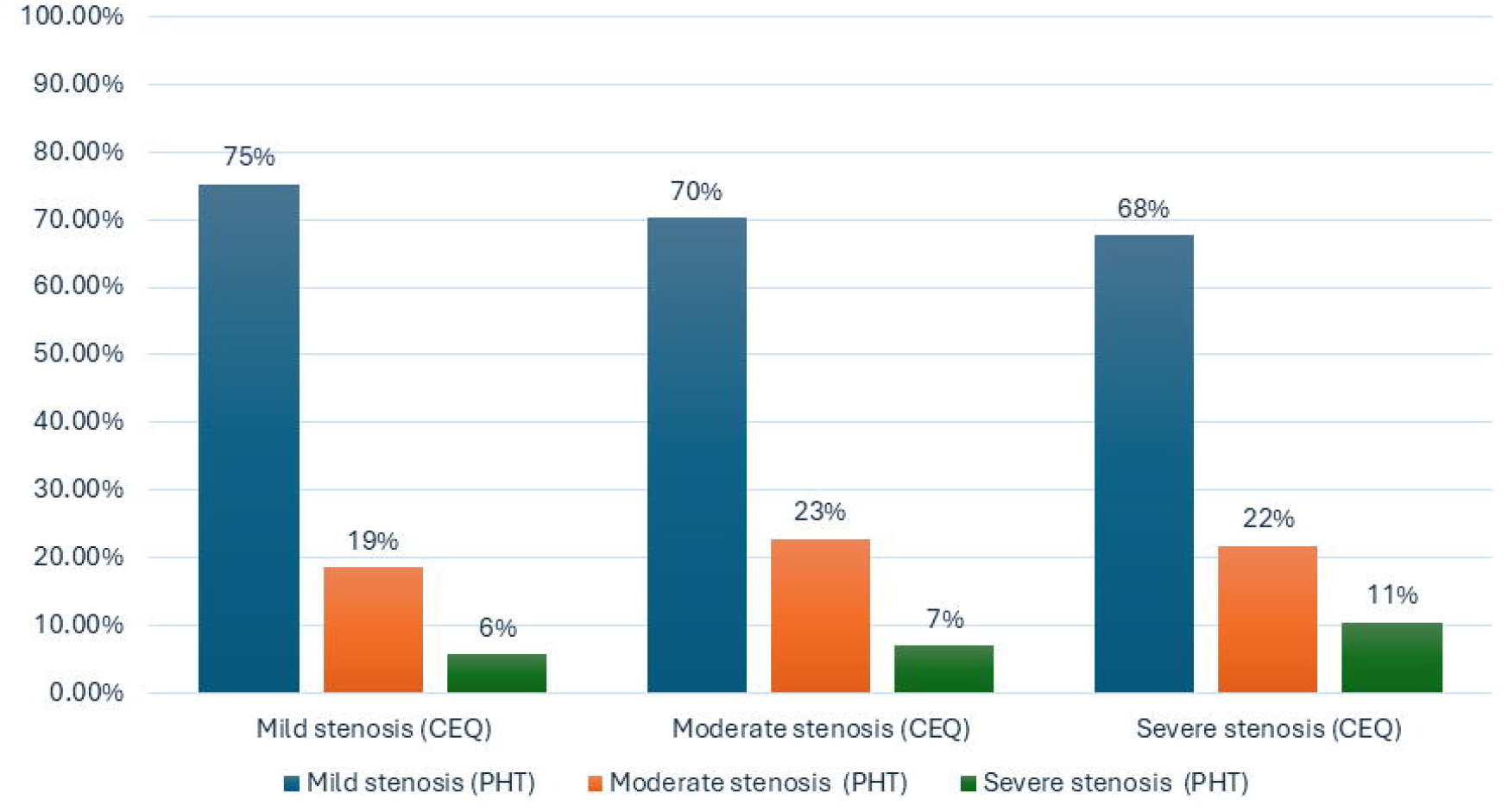
Association of mitral valve area derived by continuity equation (MVAceq) and by pressure half time (PHT)

Mean TMG was a better predictor of severe CMS with an AUC 0.66. Mean gradient >5 mm Hg had 81% specificity and 57% sensitivity for severe CMS. However, only 45% of patients with severe CMS had a mean TMG ≥ 5 mm Hg and 22% of patients with mild CMS had a mean TMG ≥ 5 mmHg. We conducted a sensitivity analysis after restricting the analysis to patients with heart rates between 60-100 beats/m(n=608) and found similar results (Table 3). We further conducted a subgroup analysis stratified by indexed stroke volume (SVi) and found that only 53% of patients with severe CMS and preserved stroke volume had mean TMG ≥ 5 mm Hg (Table 3). In patients with non-severe CMS, there was no difference between mean TMG in patients with and without preserved SVi. However, in patients with severe CMS, the mean TMG was significantly greater in patients with preserved SVi [5.3 mmHg (IQR 4-8.4)] as compared to those with reduced SVi [4.5 mmHg (IQR 3.4-6)] (p=0.01). (Figure 2)

**Table 3.**

|  | MVA<sub>cont</sub> > 2cm<sup>2</sup> | MVA<sub>cont</sub> 1.6-2 cm<sup>2</sup> | MVA<sub>cont</sub> ≤1.5 cm<sup>2</sup> |
| --- | --- | --- | --- |
| ***Overall cohort (n=717)*** |  |  |  |
| Mean TMG ≥ 5 mmHg | 72 (21.6%) | 54(22.1%) | 63(45%) |
| ***After restricting analysis to heart rates between 60-100 (n=608)*** |  |  |  |
| Mean TMG ≥ 5 mmHg | 57 (19.9%) | 45(21.2%) | 49(44.6%) |
| ***Patients with SVi ≤ 35ml/m<sup>2</sup>(n=250)*** |  |  |  |
| Mean TMG ≥ 5 mmHg | 11 (15.3%) | 18 (19.6%) | 34(39.5%) |
| ***Patients with SVi> 35ml/m<sup>2</sup>(n=457)*** |  |  |  |
| Mean TMG ≥ 5 mmHg | 61 (23.8%) | 36(24.0%) | 27 (52.9%) |

Abbreviations: mean TMG: mean transmitral gradient; SVi: indexed stroke volume

**Figure 2:**
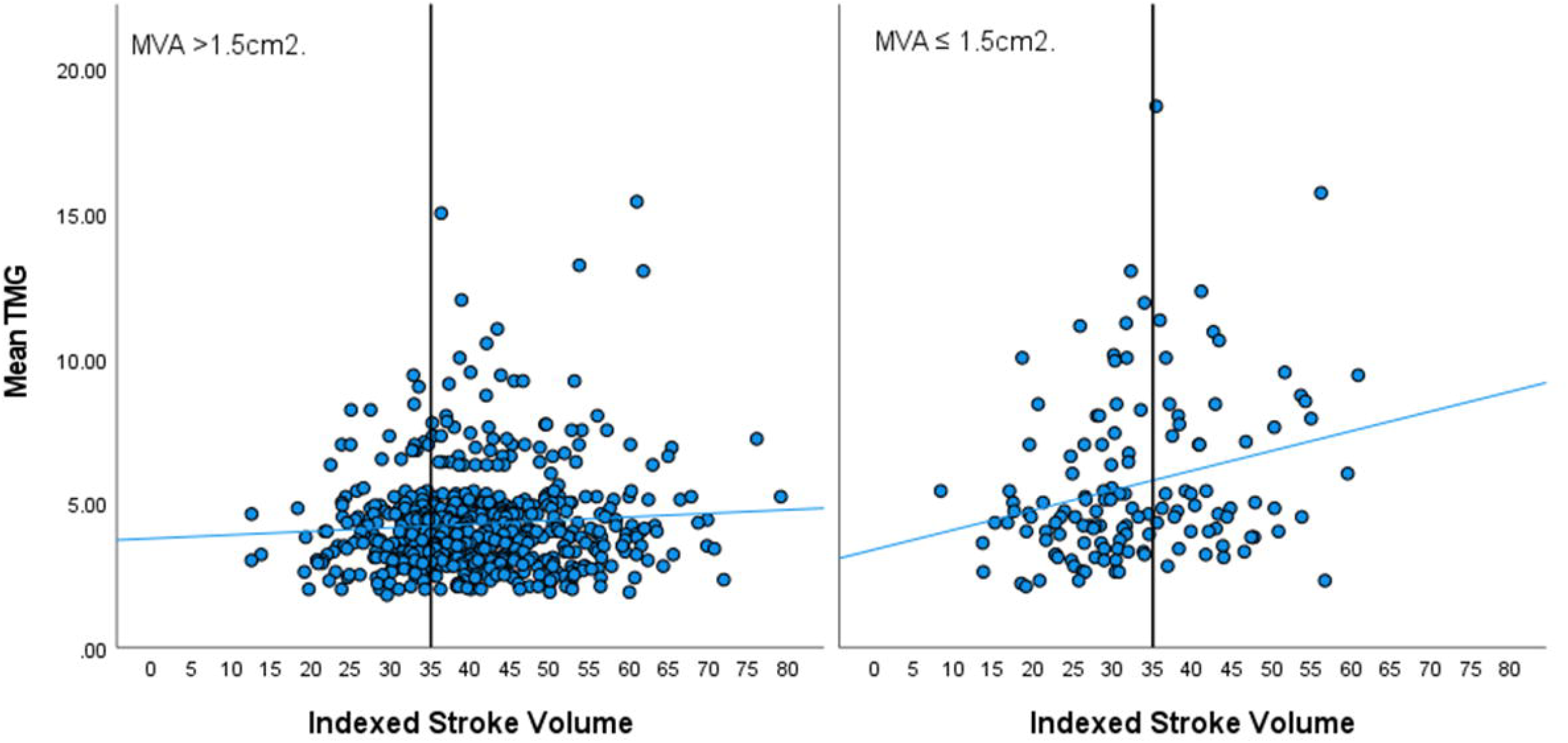
Scatterplot demonstrating association of mean TMG with indexed stroke volume in patients with and without severe calcific mitral stenosis.

### Clinical Outcomes

Over a median follow up of 36 (IQR 10.5-49.7) months, a total of n=331 (46.2%) patients died. The primary composite outcome occurred in n=370 (51.6%) patients and the secondary composite outcome occurred in n=335 patients (46.7%). Severe CMS (MVA_cont_ ≤1.5 cm^2)^ was not associated with a higher risk of either the primary outcome [aHR 1.3 (95% CI 0.9-1.8),p=0.29] or secondary outcome [aHR1.3 (95% CI 0.9-1.8), p=0.23] (Figure3). Statistically significant predictors of the primary outcome are listed in Table 4. When we replaced MVA_cont_ with mTMG in the multivariable cox-regression model, we found that a mean TMG ≥ 5 mmHg was a significant predictor of the primary outcome [aHR 1.5 (95% CI 1.1-2), p<0.01] and the secondary outcome [1.5 (95% CI 1.1-2),p=.01] (Figure 4). We further explored the data by dividing our cohort into 4 groups based on mean TMG <5 or ≥ 5 mmHg and MVA_cont_ ≤1.5 or >1.5. On multivariable cox-regression analyses using these four groups, we found that patients with MVA_cont_ ≤1.5 cm^2^ and mean gradient ≥ 5 mmHg had a significantly higher independent risk for the primary outcome [aHR 2 (95% CI 1.1-3.7),p=.02] as compared to the reference group (*patients with mean TMG <5 mmHg and MVA*_*cont*_ *>1*.*5 cm*^*2*^). The other 2 groups (MVA >1.5 cm^2^ *and* mean TMG ≥ 5 mmHg; MVA_cont_ ≤1.5 cm^2^ *and* mean TMG <5 mmHg) were not statistically different from the reference group. Similar results were seen for the secondary outcome. Patients with MVA_cont_ ≤1.5 cm^2^ and mean gradient ≥ 5 mmHg had a significantly higher independent risk for the secondary outcome [aHR 2.1 (95% CI 1.1-3.8),p=.02] as compared to the reference group. The other 2 groups were not statistically different from the reference group (Figure 5)

**Figure 3:**
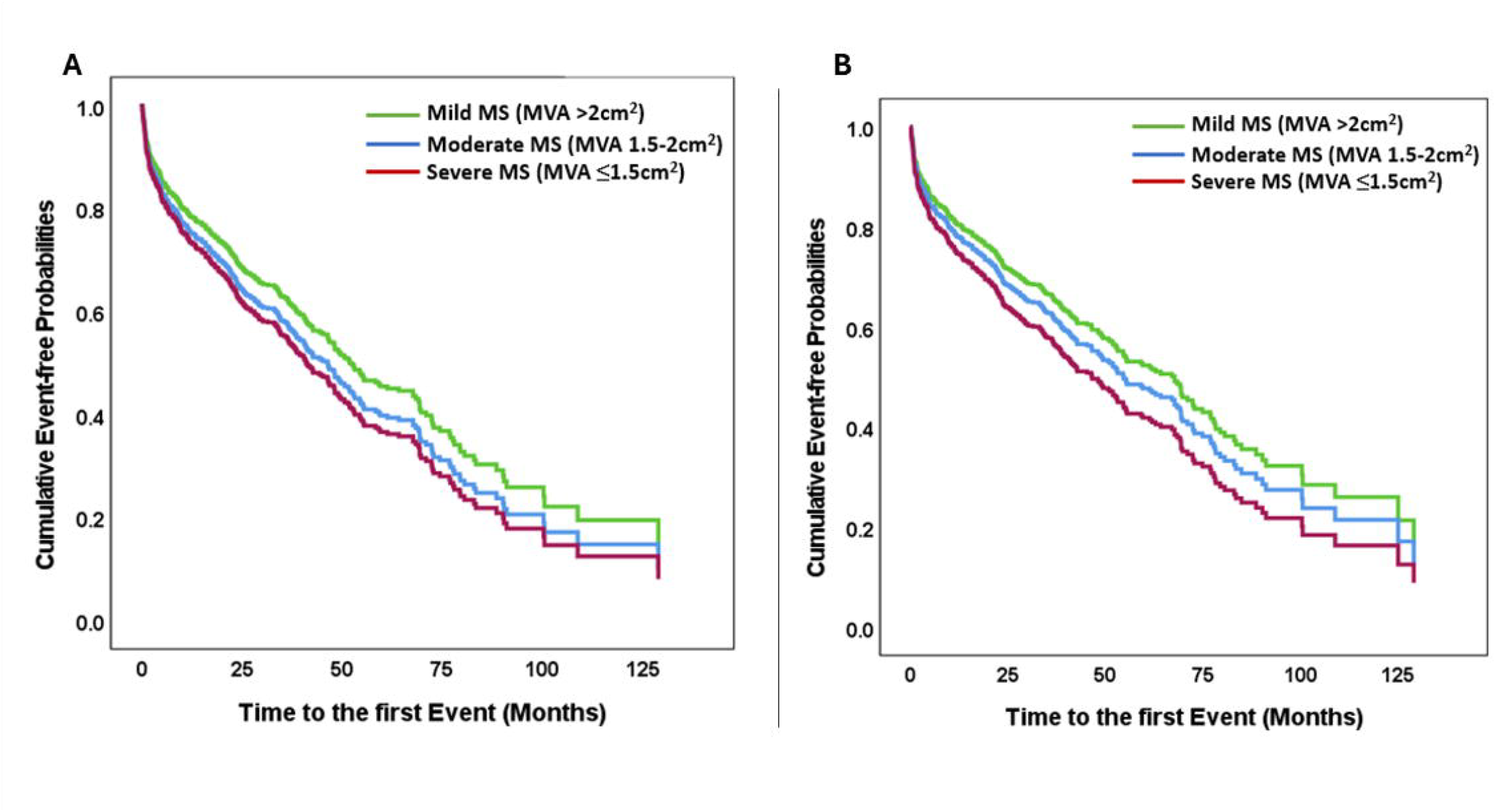
Adjusted cox-regression model demonstrating association of mild, moderate and severe CMS with the A) the primary outcome (death or mitral valve replacement or ischemic stroke) *and* B) secondary outcome (death or mitral valve replacement)

**Table 4:**
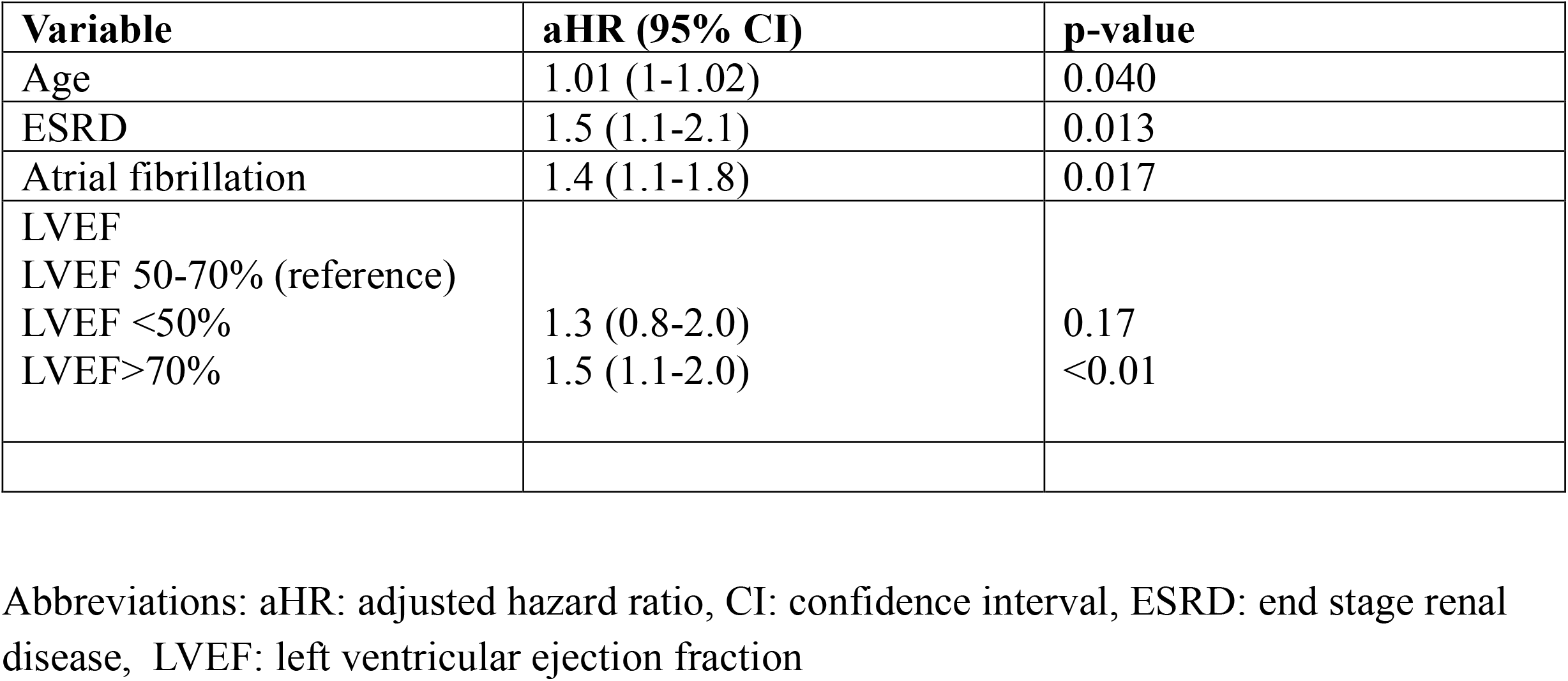
Statistically significant predictors of the primary outcome.

**Figure 4:**
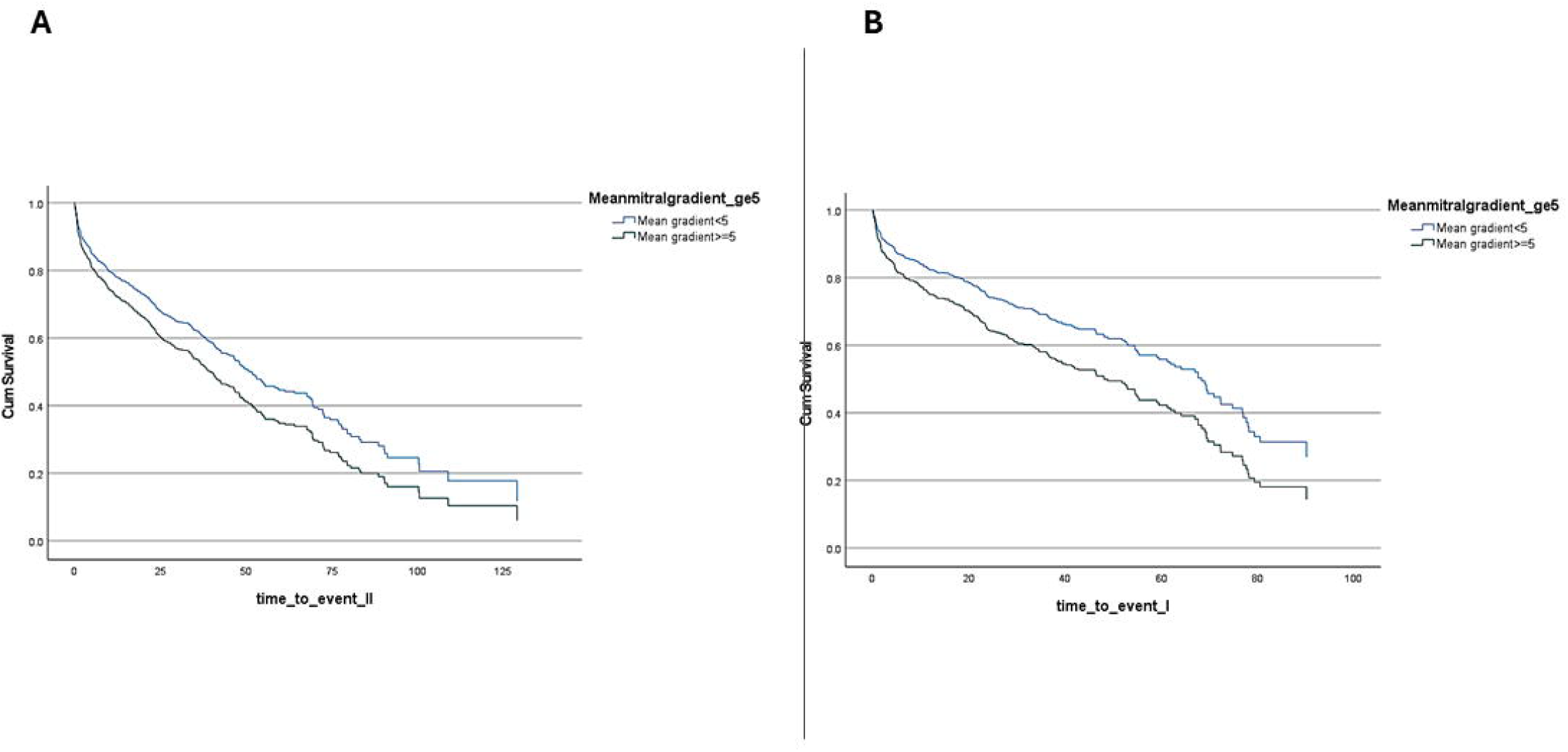
Adjusted cox-regression model demonstrating association of mean transmitral gradient groups (< 5mmHg and ≥5 mmHg) with the A) the primary outcome (death or mitral valve replacement or ischemic stroke) *and* B) secondary outcome (death or mitral valve replacement)

**Figure 5:**
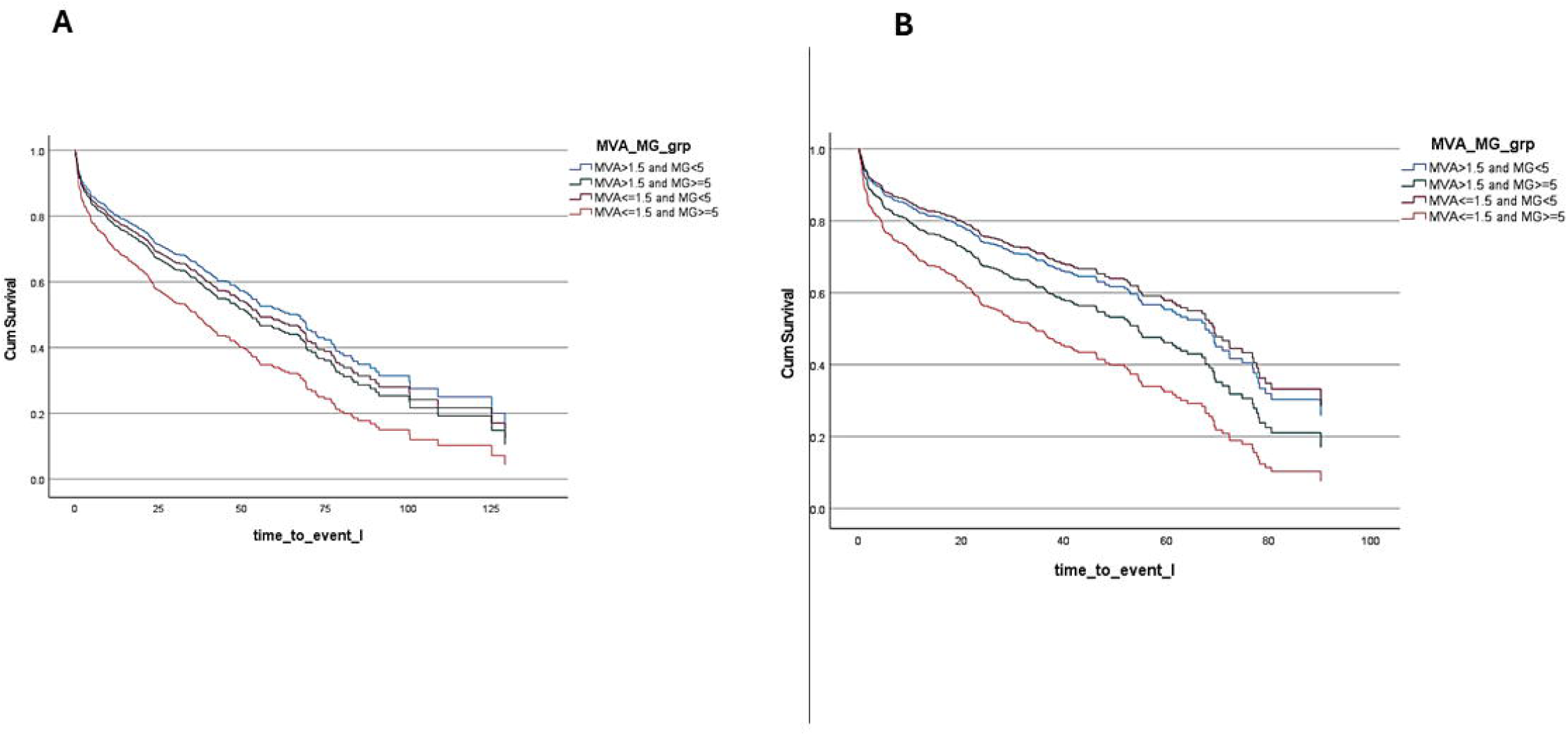
Adjusted cox-regression model demonstrating association of a combination of mitral valve area and mean transmitral groups with the A) the primary outcome (death or mitral valve replacement or ischemic stroke) *and* B) secondary outcome (death or mitral valve replacement)

## DISCUSSION

In our study of n=717 patients with CMS, we make several important observations. First, the patients are older, female, and with a high burden of comorbidities. Second, patients with severe CMS demonstrate a higher prevalence of circumferential MAC, bi-leaflet calcification and sub-valvular calcification. They also are more likely to have smaller left ventricular sizes, reduced stroke volume and right ventricular enlargement. Third, we found that overall outcomes are poor in this cohort with the primary outcome occurring in 52% of patients over a median follow-up of 3 years. Lastly, we found that mTMG (*rather than MVA*_*cont*_ *≤1*.*5 cm*^*2*^) was an independent predictor of outcomes in these patients.

MAC and CMS are highly prevalent in the older populations and in patients with renal disease, prior radiation therapy and in those with conditions which increase left ventricular pressures such as hypertrophic cardiomyopathy, hypertension and aortic stenosis.^4,12^ The pathophysiology of CMS is thought to be similar to atherosclerosis with chronic valvular stress resulting in endothelial disruption and invasion of inflammatory cells. This process leads to activation and differentiation of myofibroblasts into osteoblast-type cells, which ultimately results in calcification of the annulus and valve. ^7,13^ Our cohort had a high prevalence of atherosclerotic risk factors with 2/3rds of the patients having CAD. Of note, we found a very high proportion of women in our cohort. Our results are concordant with a prior echocardiographic study from Mayo Clinic which demonstrated that amongst those with MAC, women have 2.4-fold higher odds of developing mitral stenosis. In another natural history study, 82% of the patients with severe CMS were women.^2,10^ It is worth noting that similar trends have been reported for rheumatic heart disease with a high female to male ratio, although the mechanism for this gender disparity remains unknown.^14^

We report a poor prognosis in these patients with the primary composite outcome occurring in 52% of patients over a median follow up of 3 years. Although we included stroke as part of this composite outcome given the high prevalence of AF in this cohort as well as prior data suggesting a strong association of MAC with ischemic stroke, we found that the outcomes are primarily driven by mortality (46% during follow up).^15^

Historically, mTMG has been used as a marker of mitral stenosis severity due to its reliability and ease of measurement as well as its known prognostic value. However, it is also well recognized that mTMG can vary with hemodynamic conditions such as heart rate and cardiac output. ^16^ Despite these limitations, the 2020 American College of Cardiology/American Heart Association guidelines mention that severe mitral stenosis (valve area ≤1.5 cm2) should typically correspond to a mTMG >5 mm Hg to 10 mm Hg. In our study, we found that only 45% of patients with severe CMS demonstrate a mTMG≥ 5 mmHg, with consistent results on sensitivity analysis restricted to heart rates between 60-100 bpm as well as on subgroup analyses by indexed stroke volume. Furthermore, we found that 22% of patients with non-severe CMS have a mTMG≥ 5 mmHg. This may be explained by the fact that patients with CMS often have multiple comorbidities such as diabetes, hypertension, and CAD-all of which are associated with LV diastolic dysfunction as well as poor left atrial compliance.

These conditions can result in high filling pressures and therefore a high mTMG, which is further exaggerated by the presence of MAC and CMS-resulting in a high mTMG despite non hemodynamically significant stenosis.^4,5^ However, despite these pit-falls, we found mTMG (*rather than MVA*) to be an independent predictor of outcomes in CMS. This is a critical finding and suggests that patients with MVA ≤1.5 cm^2^ who do not have gradients >5 mmHg, may have a better prognosis and perhaps may benefit from deferral of valvular intervention. This is especially important in light of the fact that valvular interventions for CMS, both surgical and percutaneous-are challenging and associated with poor procedural and long-term outcomes.^7,17–21^ Our findings are concordant with the single center study by Kato *et al* which showed that a mTMG ≥8 mm Hg had independent prognostic value in patients with severe CMS. Although further data are needed, we suggest that future trials to evaluate valvular interventions in CMS, consider the use of mTMG in addition to MVA as an inclusion criteria given its prognostic significance. Of note, another interesting aspect of our analysis was the association of hyperdynamic left ventricular function with poorer outcomes. These findings are in line with a prior study demonstrating increased mortality in patients with supranormal LVEF, especially amongst women, those with concentric remodeling and with reduced stroke volume.^22^ We postulate that in our cohort of predominantly older women, presence of hypertrophied ventricles with smaller end-diastolic volumes possibly contributes to lower stroke volumes and therefore a compensatory increase in LVEF. Further research is needed to reproduce and validate these results.

Our study has several limitations. First, this is a single center, retrospective study where echocardiograms were performed for varied indications including patients hospitalized with other cardiovascular conditions. This generates a selection bias with the cohort possibly having a higher burden of comorbidities (and therefore poorer outcomes) than the general population of patients with CMS. Second, we were unable to use right ventricular systolic pressure in our cox-regression model due to a high percentage of missing values. Lastly, as the continuity equation was used for evaluation of MVA, a minority of patients were excluded due to inability to measure the LVOTd. Despite these limitations, our study provides comprehensive echocardiographic data on CMS and adds to the limited literature on natural history and prognostic makers.

## CONCLUSION

Patients with severe CMS are older, female with a high burden of comorbidities and carry an overall poor prognosis. mTMG is an independent prognostic marker in these patients. Patients with MVA ≤1.5 cm^2^ and mTMG ≥5 mmHg have the worst prognosis.

## Data Availability

Data are not available upon request

## ACKNOWLEDGEMENTS

This research was supported by an internal grant from the Cardiovascular Academic Initiative (Division of Cardiology, Medical College of Wisconsin)

