## Supplement for "Echocardiographic Characteristics, Measures of Severity and Natural History of Isolated Calcific Mitral Stenosis"

Figure 1

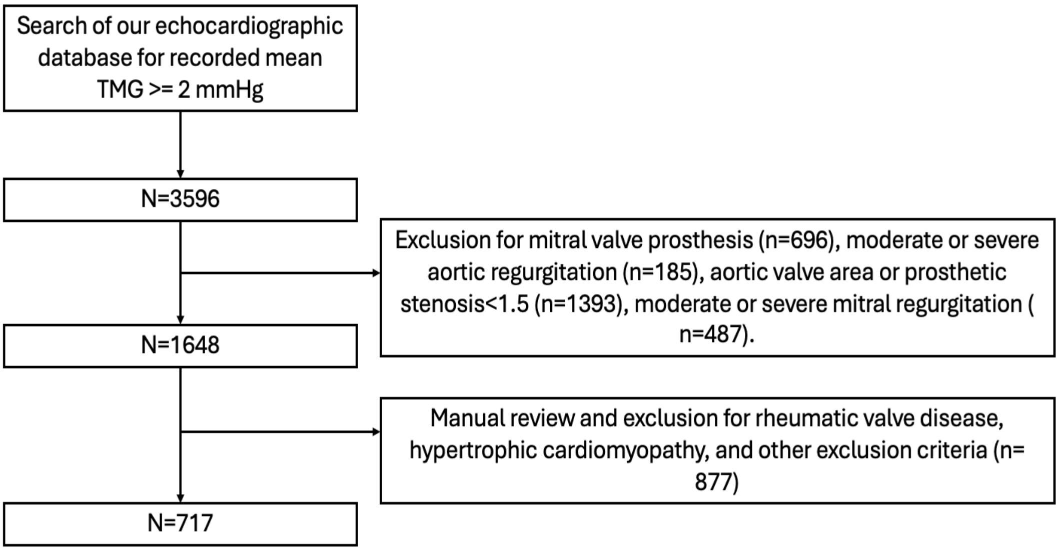


Figure 2 ROC analysis of $MVA_{cont}$ and $MVA_{PHT}$

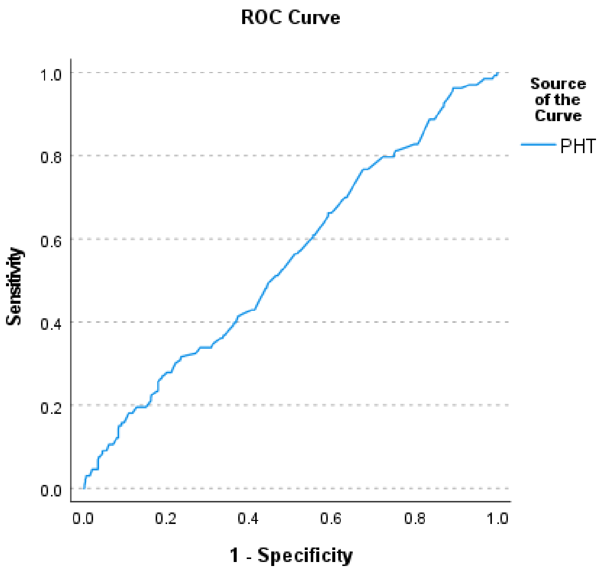


Figure 3: ROC analysis of $MVA_{cont}$ and mean transmitral gradient.

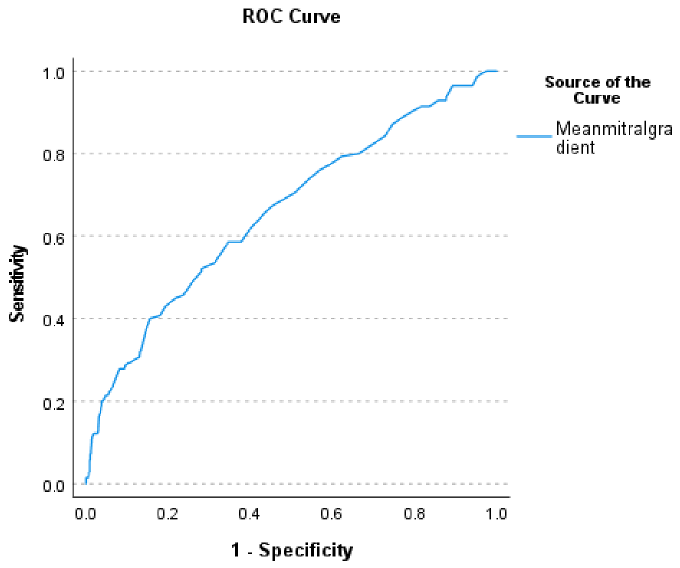
ROC Curve
Sensitivity
1.0
0.8
0.6
0.4
0.2
0.0
0.0
0.2
0.4
0.6
0.8
1.0
1 - Specificity
Source of the Curve
Meanmitralgra dient
